# Association of dietary protein intake with accelerated biological aging markers and all-cause mortality in adults

**DOI:** 10.64898/2026.10.27.26366537

**Authors:** Juan Reyes-Barrera, Omar Yaxmehen Bello-Chavolla, Carlos A. Fermín-Martínez, Ashuin Kammar-García, Nestor Alonso Sánchez-Ortiz, Rogelio Reyes Reyes, Fernando Enrique García Arroyo, Neftali Eduardo Antonio-Villa

**Author notes:** Lead authorship (Juan Reyes-Barrera). Last authorship (Neftali Eduardo Antonio-Villa). **Correspondence: a)** Juan Reyes-Barrera. Departamento de Endocrinología, Instituto Nacional de Cardiología Ignacio Chávez, Juan Badiano 1, Belisario Domínguez Secc 16, Tlalpan, 14080, Mexico City, Mexico., **b)** Neftali Eduardo Antonio-Villa. Departamento de Endocrinología, Instituto Nacional de Cardiología Ignacio Chávez, Juan Badiano 1, Belisario Domínguez Secc 16, Tlalpan, 14080, Mexico City, Mexico. **Declaration of Competing Interest:** Nothing to disclose.

## Abstract

**BACKGROUND:** Aging processes could be influenced by diet. However, the extent to which dietary protein intake is associated with biological aging (BA) markers remains poorly characterized.

**OBJECTIVES:** Here, we evaluated the association between dietary protein intake and BA markers and explored the extent to which BA markers mediate the association between protein intake and all-cause mortality.

**METHODS:** We analyzed data from NHANES cycles from 1999-2000 and 2001-2002. Dietary protein intake was expressed in g/kg/day. BA was estimated with TL, AnthropoAge, PhenoAge, and the Klemera–Doubal Method (KDM). Accelerated aging was defined as TL<25^th^ percentile of the weighted-population distribution and as residuals from BA markers regressed against CA. Survey-weighted logistic and linear regression models were fitted. Mortality correlates were evaluated using cause-specific Cox proportional hazards models and mediation analyses.

**RESULTS:** Among 6,249 adults (median age: 45 [IQR: 34–58], 52% women), we found that participants in the highest protein intake quintile (≥1.40 g/kg/day) had a lower adjusted likelihood of having accelerated aging by TL (OR 0.40, 95% CI: 0.22-0.73), PhenoAge (OR: 0.54, 95% CI: 0.32-0.92), and KDM (OR: 0.53, 95% CI: 0.32-0.87), compared to those in lowest quintile (≤0.62 g/kg/day). Higher dietary protein intake was associated with a lower hazard of all-cause mortality, and BA metrics partially mediated these associations.

**CONCLUSIONS:** Higher dietary protein intake was associated with lower likelihood of accelerated aging and reduced mortality, with the latter association partially mediated by BA markers. Our findings suggest that dietary protein intake could influence healthy aging.

## INTRODUCTION

Aging is characterized by a progressive accumulation of molecular and physiological changes that lead to functional decline, chronic disease, and increased mortality (1). However, one of the primary challenges in Geroscience is the lack of consensus on the definition of aging. An approach proposed by Salin and Levine *et al.* posits that the divergence between chronological age (CA) and accumulation of biological changes can be reflected in an individual’s biological age (BA) (2). Therefore, some BA proxies have been proposed to capture molecular, cellular, and clinical age-related changes and their underlying mechanisms (3). However, given the multifactorial nature of aging, it is unlikely that a single marker can fully capture all physiological domains affected by aging (4). Accordingly, numerous BA candidates have been proposed, although no single “gold-standard” marker has yet been fully established (5).

Among molecular markers, telomere length (TL) progressively shortens with each cell division (6). Although some studies have highlighted limitations related to variability, lack of standardization, and constraints on tissue sampling, it remains the most widely used BA marker in epidemiological studies (6). Consequently, there has been growing interest in other BA markers that capture different domains of aging. The Klemera-Doubal Method (KDM), for instance, is a first-generation BA clock that combines chronological age (CA) with weighted clinical biomarkers according to their statistical association with CA (7). Other BA markers, such as Phenotypic Age (PhenoAge) and Anthropometric Age (AnthropoAge), represent second-generation BA clocks that estimate mortality risk using combinations of blood biomarkers and anthropometric measures (8). Therefore, TL and these three BA markers likely provide complementary information across different BA domains and can be used to examine potential contributors.

Diet, particularly dietary protein intake, has emerged as a potential modifiable contributor of BA (9). Dietary protein is essential for muscle maintenance, immune function, and metabolic regulation, all of which can modulate an individual’s aging process (9). Although dietary protein intake has been explored as a potential contributor to BA, these studies have not examined the association with novel BA metrics, such as AnthropoAge, nor have they assessed whether these markers mediate associations between dietary protein intake and mortality (10). In this study, we aim 1) to evaluate the association between dietary protein intake and BA markers and 2) to evaluate the association of protein intake and explore the mediating effect of BA in the relationship with all-cause and cause-specific mortality using data from participants enrolled in the National Health and Nutrition Examination Surveys (NHANES).

## METHODS

### Study population

This study is a sub-analysis of NHANES.. Detailed sampling and assessment methods have been published elsewhere (11). Briefly, NHANES comprises a series of cross-sectional surveys conducted in the United States since the 1960s by the National Center for Health Statistics (NCHS) and endorsed by the Centers for Disease Control and Prevention. NHANES uses a complex, multistage, probability sampling design to obtain a nationally representative sample of the U.S. population. NHANES has received approval from the NCHS Ethics Committee. For this sub-analysis, we analyzed data of adults ≥20 years from the 1999–2000 and 2001–2002 cycles with available dietary data and leukocyte TL. Among 21,004 participants who underwent examination and consented to DNA analysis, we excluded individuals with missing dietary, TL, or BA-related data (12). Because NHANES data are publicly available, additional local ethics approval was not required. All participants provided informed consent. This study follows the Strengthening the Reporting of Observational Studies in Epidemiology (STROBE) guidelines (**Supplementary Table 1**).

### Exposure assessment

The main exposure was daily protein intake (g/kg/day), as used in previous studies (13). Dietary assessment methods are summarized in **Supplementary Table 2**. Briefly, dietary intake was assessed using 24-hour recalls. Protein intake included animal (meat, poultry, seafood, dairy, and eggs) and plant (legumes, nuts, seeds, and soy products) sources (14). Daily protein intake (g/kg/day) was calculated by dividing total protein intake (g/day) by body weight (13,15). Weighted-population quintiles were defined as follows: Q1, <0.62; Q2, 0.62–0.85; Q3, 0.85–1.07; Q4, 1.07–1.40; and Q5, >1.40 g/kg/day. Quintiles were derived using NHANES sampling weights. Protein intake recommendations were based on the recommended dietary allowance (RDA) of 0.8 g/kg/day (16). Dietary protein intake and BA markers were assessed during the same examination visit.

### Outcomes definitions

This study focused on two primary outcomes: 1) BA markers and 2) all-cause and cause-specific mortality.

#### a) Biological aging markers

TL was used as the primary marker of BA.Methods for TL assessment are described in Supplementary Table 3. To define TL-based accelerated aging (TLAgeAccel), participants with TL values below the 25th percentile (<0.84) were classified as having accelerated aging, whereas those with values ≥0.84 were considered to have normative aging. (18–20). Three complementary BA markers were also evaluated: AnthropoAge, PhenoAge, and the Klemera-Doubal Method (KDM) (8,20,21). Their estimation methods and included variables are summarized in Supplementary Table 4. Briefly, AnthropoAge was calculated using chronological age (CA), BMI, and waist-to-height ratio (WHtR) (22,23). PhenoAge was calculated based on nine blood biomarkers (albumin, alkaline phosphatase, creatinine, C-reactive protein, glucose, mean cell volume, red cell distribution width, white blood cell count, and lymphocyte proportion) plus CA (24). KDM was calculated from a set of regressions based on biomarkers of CA and represents the age at which the mean physiological function matches the individual’s physiological function. The KDM approach included the following variables: forced expiratory volume in one second (FEV_1_), systolic blood pressure, and seven blood biochemical variables (albumin, alkaline phosphatase, blood urea nitrogen, creatinine, C-reactive protein, glycosylated hemoglobin, and total cholesterol) (25). BA acceleration of these metrics was assessed by computing residuals from linear regression of BA onto CA. The resulting residuals, termed AnthropoAgeAccel, PhenoAgeAccel, and KDMAgeAccel reflect the deviation of BA from CA. Residuals >0 indicate accelerated aging, values ≤0 indicate non-accelerated or normative aging.

#### b) All-cause and cause-specific mortality

Mortality outcomes were obtained from the NHANES Linked Mortality Files. Follow-up was calculated from the NHANES examination date until death or December 31, 2019, whichever occurred first; surviving participants were censored. Mortality status and causes of death were identified using International Classification of Diseases, 10th Revision (ICD-10) codes. Cause-specific mortality included heart disease, malignant neoplasms, and cerebrovascular disease..

### Covariate assessment

NHANES participants completed standardized questionnaires collecting demographic data, including sex, age, poverty-to-income ratio (PIR), and ethnicity (Caucasian, African

American, Mexican American, or other). Lifestyle variables included physical activity (moderate- or vigorous-intensity activity) and current smoking. Self-reported chronic conditions included type 2 diabetes (T2D) and hypertension. Anthropometric measurements included height, BMI, waist circumference, and blood pressure. Overweight and obesity were defined as BMI ≥25 and ≥30 kg/m², respectively. Central obesity was defined as waist circumference ≥102 cm in men and ≥88 cm in women (26). Biochemical variables included plasma triglycerides, total cholesterol, high-density lipoprotein cholesterol (HDL-C), and glucose. Hypertriglyceridemia was defined as triglyceride levels ≥150 mg/dL, and low HDL-C was defined as HDL-C levels <40 mg/dL in men and <50 mg/dL in women. Dietary covariates included total energy intake, energy from carbohydrates, and energy from fat (Kcal/day). All covariates were self-reported or measured during the NHANES examination and incorporated into the survey-weighted models.

#### Statistical analysis

To account for the NHANES design, survey weights were applied to obtain nationally representative estimates. Specifically, 4-year sample weights (WTMEC4YR), strata (SDMVSTRA), and cluster variables (SDMVPSU) were used as recommended by NHANES analytic guidelines for pooled dataAnalyses were conducted using the R survey package(28). Code is publicly available on GitHub at https://github.com/REYJUA12235/Protein-and-Aging. Statistical analyses were performed in R (version 4.3.2), and p < 0.05 was considered statistically significant

### Association between protein intake, aging markers, and mortality outcomes

First, we evaluated correlations between TL and other aging markers using Spearman coefficients. To assess associations between dietary protein intake and BA markers, we performed linear regression analyses using protein intake as both a continuous variable (g/kg/day) and quintiles. Outcomes included TL (T/S ratio), AnthropoAge, PhenoAge, and KDM. We then fitted binomial logistic regression models to estimate odds ratios (ORs) for accelerated aging (TLAgeAccel, AnthropoAgeAccel, PhenoAgeAccel, and KDMAgeAccel) according to protein intake (continuous and quintiles; Q1 as reference). Confounders were selected using a direct acyclic graph (DAG) following *Tennant et al.* (29). According to Supplementary Figure 1, adjusted models included age-residualized BA markers, sex, total energy intake, ethnicity, PIR, smoking, physical activity, hypertension, and T2D.

To support these findings, two sensitivity analyses were performed. First, age-stratified linear regression models were conducted in participants aged <40, 40–60, and >60 years, including interaction terms between protein intake and age category (<40 years as reference). Second, total energy intake was replaced by carbohydrate intake (Model 1) or fat intake (Model 2), while retaining the remaining covariates.

Finally, associations between protein intake, BA markers, and all-cause and cause-specific mortality were evaluated using cause-specific Cox proportional hazards models, reporting adjusted hazard ratios (aHRs). To explore mediation, joint models including protein intake and each BA marker were fitted simultaneously. Analyses were restricted to complete cases; therefore, no imputation was performed.

### Mediation analysis

To explore whether BA markers mediate the association between protein intake (E) and all-cause or cause-specific mortality (Y), we performed survey-weighted mediation analyses using a regression-based framework following *Zhang et al.* (30). Separate models were fitted for each BA marker (M). Models estimated direct and indirect effects of protein intake on mortality outcomes. All models were adjusted for the same confounders used in previous analyses.

## RESULTS

### Study population

We included data from 6,249 participants for this sub-analysis (**Supplementary Figure 2**). The median CA of the study population was 45 (IQR: 34–58), with 51.7% of participants being women and 82.6% Caucasian. The median TL was 0.99 (IQR: 0.84–1.17), whereas the median AnthropoAge was 45.3 years (IQR: 34.6–59.2), PhenoAge was 41 years (IQR: 30.1–54.9), and KDM was 24.4 years (IQR: 17.8–33.1). **Table 1** summarizes participant characteristics by dietary protein intake quintile. Participants in the highest quintile were younger, more often male, predominantly Caucasian, and had a higher PIR and total energy intake than those in the lowest quintile. Physical activity did not differ across quintiles. Individuals in the lowest quintile had higher BMI and waist circumference, greater prevalence of obesity, central obesity, T2D, and hypertension, and a less favorable biochemical profile. Regarding BA markers, higher protein intake quintiles were associated with higher TL and lower AnthropoAge, PhenoAge, and KDM values. The prevalence of accelerated aging also declined across increasing protein intake quintiles. Dietary intake characteristics are presented in **Supplementary Table 5**. Median protein intake was 0.95 g/kg/day (IQR: 0.68–1.30) and 1.09 g/kg/day (IQR: 0.80–1.45) when adjusted for ideal body weight. Overall, 62% of the weighted sample met the recommended dietary allowance for protein.

**Table 1:** Characteristics of study participants according to quintiles of protein intake (g/kg/d).

|  | Protein intake quintiles (g/kg/d) |  |  |  |  | *p-value* |
| --- | --- | --- | --- | --- | --- | --- |
|  | Q1, n = 1,353 | Q2, n = 1,313 | Q3, n = 1,233 | Q4, n = 1,189 | Q5, n = 1,161 |  |
|  | (<0.62) | (0.62–0.85) | (0.85–1.07) | (1.07–1.40) | (>1.40) |  |
| ***Sociodemographic*** |  |  |  |  |  |  |
| Age (years) | 47.0 (37.0, 67.0) | 52.5 (38.0, 67.0) | 50.0 (36.0, 67.0) | 47.0 (34.0, 64.0) | 41.0 (30.0, 55.0) | <0.001 |
| Sex, female, n (%) | 839 (63.8) | 740 (57.1) | 617 (48.9) | 551 (46.4) | 499 (42.2) | <0.001 |
| Caucasic, n (%) | 662 (78.0) | 749 (82.6) | 729 (84.8) | 726 (85.8) | 643 (82.0) | <0.001 |
| Afro-American, n (%) | 363 (15.5) | 234 (9.9) | 171 (7.4) | 162 (7.5) | 178 (8.6) | <0.001 |
| Mexican American, n (%) | 328 (6.6) | 330 (7.5) | 333 (7.8) | 301 (6.8) | 340 (9.4) | <0.001 |
| Poverty to income ratio | 2.5 (1.3, 4.5) | 3.4 (1.8, 5.0) | 3.4 (1.8, 5.0) | 3.5 (1.8, 5.0) | 3.5 (1.8, 5.0) | <0.001 |
| ***Lifestyle habits*** |  |  |  |  |  |  |
| Active smoker, n (%) | 298 (27.0) | 258 (22.7) | 233 (20.6) | 255 (23.7) | 294 (26.1) | 0.009 |
| Moderate physical activity, n (%) * | 515 (80.1) | 563 (76.9) | 545 (77.9) | 538 (77.2) | 561 (78.0) | 0.8 |
| Energy (kcal) | 1,289.2 (982.5, 1,671.0) | 1,739.2 (1,418.0, 2,210.8) | 2,077.0 (1,692.9, 2,595.4) | 2,370.0 (1,926.5, 2,982.2) | 2,983.0 (2,336.0, 3,713.6) | <0.001 |
| ***Anthropometric measures*** |  |  |  |  |  |  |
| BMI (kg/m<sup>2</sup>) | 30.2 (26.3, 35.7) | 28.5 (24.9, 32.6) | 27.5 (24.5, 31.2) | 26.1 (22.9, 29.2) | 24.4 (21.7, 27.4) | <0.001 |
| Overweight, n (%) | 424 (30.6) | 491 (35.1) | 487 (38.8) | 468 (36.8) | 422 (34.2) | 0.019 |
| Obesity, n (%) | 711 (51.6) | 536 (39.3) | 400 (31.6) | 259 (20.9) | 130 (10.0) | <0.001 |
| WC (cm) | 102.0 (91.7, 113.6) | 98.2 (88.4, 109.4) | 97.3 (87.5, 106.7) | 93.1 (81.7, 102.6) | 88.2 (78.9, 96.2) | <0.001 |
| Central obesity, n (%) | 986 (71.8) | 848 (60.3) | 696 (53.8) | 516 (40.1) | 305 (22.5) | <0.001 |
| SBP (mmHg) | 122.0 (112.0, 134.0) | 122.0 (112.0, 132.0) | 120.0 (110.0, 134.0) | 118.0 (110.0, 130.0) | 116.0 (106.1, 126.0) | <0.001 |
| DBP (mmHg) | 74.0 (66.0, 80.0) | 74.0 (66.0, 80.0) | 72.0 (66.0, 80.0) | 74.0 (66.0, 80.0) | 72.0 (64.0, 79.6) | 0.005 |
| ***Biochemical data*** |  |  |  |  |  |  |
| Triglycerides (mg/dL) * | 121.0 (86.0, 183.0) | 119.0 (81.0, 173.0) | 123.0 (85.9, 172.0) | 109.0 (68.0, 158.0) | 104.0 (67.0, 153.0) | <0.001 |
| Total cholesterol (mg/dL) | 204.0 (180.0, 231.0) | 206.0 (180.0, 233.0) | 201.0 (179.0, 230.0) | 199.0 (177.0, 225.0) | 197.0 (171.1, 225.0) | <0.001 |
| HDL-C (mg/dL) | 47.0 (39.0, 58.0) | 49.0 (40.0, 59.0) | 48.0 (41.0, 58.0) | 50.0 (41.0, 63.0) | 51.0 (42.0, 63.0) | <0.001 |
| Glucose (mg/dL)* | 89.0 (84.0, 98.0) | 89.0 (83.0, 97.0) | 90.0 (84.0, 96.0) | 89.0 (82.0, 96.0) | 87.0 (82.0, 93.0) | 0.001 |
| ***Concomitants comorbidities*** |  |  |  |  |  |  |
| T2D, n (%) | 150 (8.0) | 138 (7.4) | 111 (6.6) | 81 (3.8) | 50 (2.9) | <0.001 |
| Arterial hypertension, n (%) | 513 (34.3) | 446 (29.2) | 396 (27.4) | 312 (24.5) | 188 (14.7) | <0.001 |
| Hypertriglyceridemia, n (%) * | 252 (35) | 243 (34) | 235 (35) | 169 (28) | 179 (27) | 0.043 |
| Low HDL-C, n (%) | 596 (46) | 497 (37) | 420 (36) | 355 (31) | 314 (27) | <0.001 |
| ***Biological age markers*** |  |  |  |  |  |  |
| Telomere length | 0.98 (0.83, 1.15) | 0.99 (0.85, 1.18) | 1.02 (0.87, 1.20) | 1.02 (0.87, 1.20) | 1.05 (0.90, 1.22) | <0.001 |
| AnthropoAge (years) | 49.3 (36.3, 64.9) | 48.3 (37.4, 62.8) | 46.0 (35.7, 60.2) | 44.6 (34.7, 55.9) | 39.9 (30.5, 51.7) | <0.001 |
|  | Q1, n = 1,353<br>(<0.62) | Q2, n = 1,313<br>(0.62–0.85) | Q3, n = 1,233<br>(0.85–1.07) | Q4, n = 1,189<br>(1.07–1.40) | Q5, n = 1,161<br>(>1.40) |  |
| PhenoAge (years) | 44.9 (31.6, 60.0) | 43.4 (33.2, 58.8) | 42.3 (31.1, 55.7) | 40.5 (30.8, 52.6) | 35.3 (25.5, 46.8) | <0.001 |
| KDM (years) | 27.9 (20.8, 36.4) | 26.7 (19.3, 35.9) | 24.6 (18.1, 33.6) | 23.2 (17.3, 31.1) | 20.2 (15.1, 27.7) | <0.001 |
| ***Accelerated aging markers*** |  |  |  |  |  |  |
| TLAgeAccel, n (%) | 394 (27.1) | 344 (22.9) | 310 (21.3) | 295 (20.7) | 219 (15.7) | <0.001 |
| AnthropoAgeAccel, n (%) | 649 (43.6) | 506 (34.6) | 445 (31.7) | 407 (31.8) | 398 (30.1) | <0.001 |
| PhenoAgeAccel, n (%) | 678 (47.6) | 567 (38.0) | 535 (38.9) | 459 (34.0) | 458 (37.1) | <0.001 |
| KDMAgeAccel, n (%) | 765 (55.6) | 662 (48.3) | 581 (41.8) | 476 (39.5) | 442 (35.2) | <0.001 |

Footnotes: Data are presented as median (IQR) for continuous variables and n (weighted %) for categorical variables. Overweight was defined as BMI $\geq 25$ kg/m<sup>2</sup> and obesity as BMI $\geq 30$ kg/m<sup>2</sup>. Central obesity was defined as waist circumference $\geq 102$ cm for men and $\geq 88$ cm for women. Hypertriglyceridemia was defined as triglyceride levels $\geq 150$ mg/dL, and low HDL-C as HDL cholesterol $< 40$ mg/dL in men and $< 50$ mg/dL in women. Accelerated aging refers to aging acceleration based on each specific metric (telomere length, PhenoAge, KDM, or AnthropoAge). AnthropoAge: Anthropometric Age; PhenoAge: Phenotypic Age; KDM: Klemmera and Doubal method for biological age estimation. P-values were obtained using survey-weighted tests. Percentages are weighted according to the NHANES complex survey design, whereas counts represent unweighted sample sizes, and protein intake quintiles were defined using survey-weighted distributions, representing equal proportions of the population rather than equal sample sizes.
Abbreviations: BMI, body mass index; WC, waist circumference; HDL-C, high-density lipoprotein cholesterol; SBP, systolic blood pressure; DBP, diastolic blood pressure; TL, telomere length.
a) Sample sizes varied across variables due to missing data: physical activity (n = 3,516), triglycerides (n = 3,216), and glucose (n = 3,216).

### Association of dietary protein intake and aging markers

TL showed moderate inverse correlations with AnthropoAge, PhenoAge, and KDM (**Supplementary Figure 3**). We then evaluated associations between dietary protein intake and BA markers (**Table 2**). Higher protein intake was associated with longer TL and lower AnthropoAge, PhenoAge, and KDM. Likewise, participants in the highest protein intake quintile had lower BA estimates than those in the lowest quintile. For every 1 g/kg/day increase in protein intake, the odds of accelerated aging were lower for TLAgeAccel, PhenoAgeAccel, and KDMAgeAccel, although the association with AnthropoAgeAccel was attenuated after adjustment. Similarly, participants in the highest quintile had lower odds of TLAgeAccel (OR 0.40, 95% CI 0.22–0.73), PhenoAgeAccel (OR 0.54, 95% CI 0.32–0.92), and KDMAgeAccel (OR 0.53, 95% CI 0.32–0.87) than those in the lowest quintile. For AnthropoAgeAccel, a significant association was observed only in the third quintile (OR 0.58, 95% CI 0.34–0.99) (Figure 1). Sensitivity analyses showed consistent associations across age groups, although TL and PhenoAge tended to show stronger associations in older (>60 years) and middle-aged (40–60 years) participants than in younger adults (<40 years) (**Supplementary Table 6**). Associations remained unchanged after adjustment for nutrient-specific energy intake (**Supplementary Table 7**).

**Figure 1.**
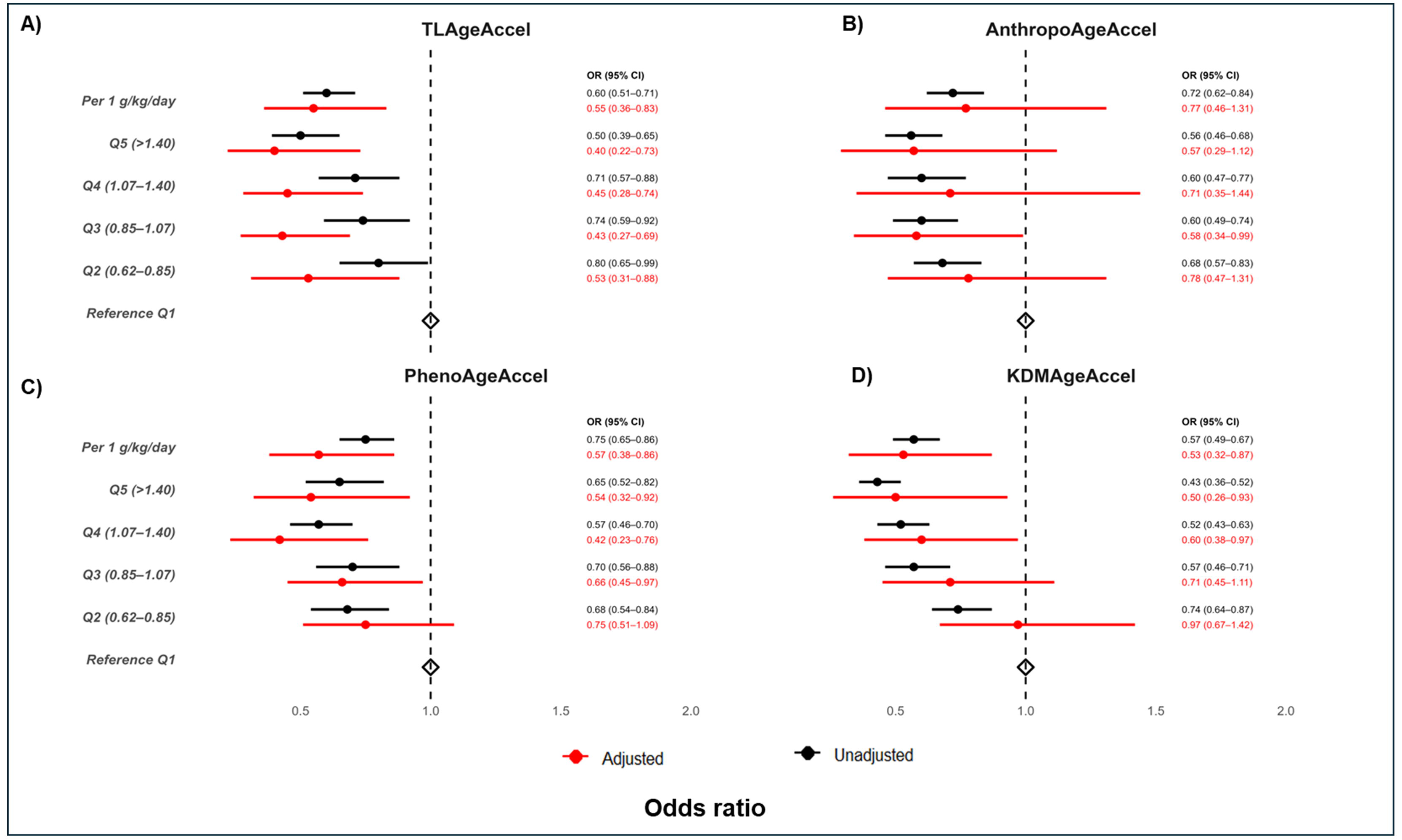
Association between protein intake and accelerated biological aging markers. The adjusted model includes age (residual), sex, energy (Kcal), ethnicity, poverty-to-income ratio, tobacco use, physical activity, hypertension, and diabetes. Accel refers to aging acceleration based on each specific metric: PhenoAge, KDM, or AnthropoAge.

**Table 2.**
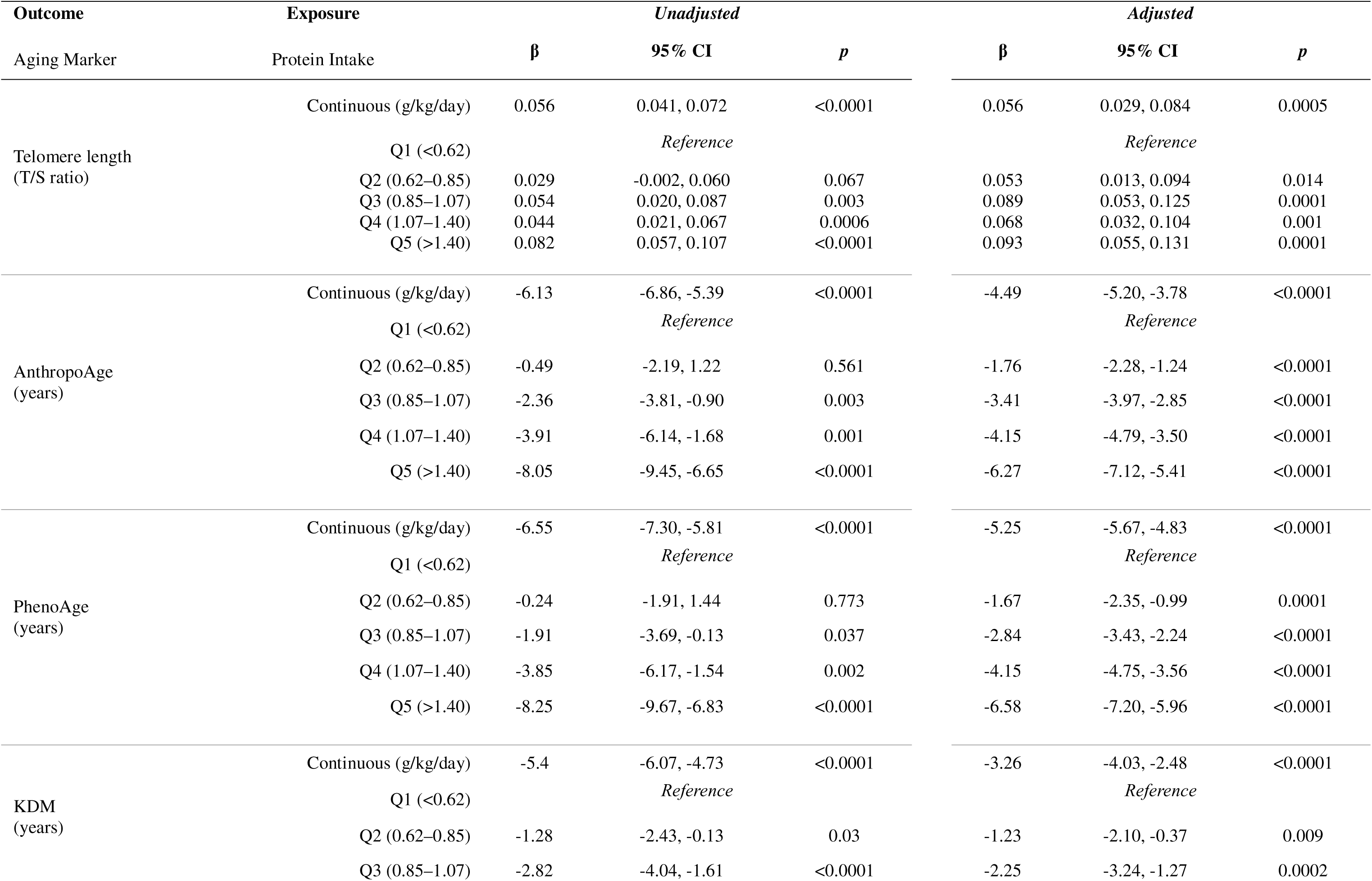
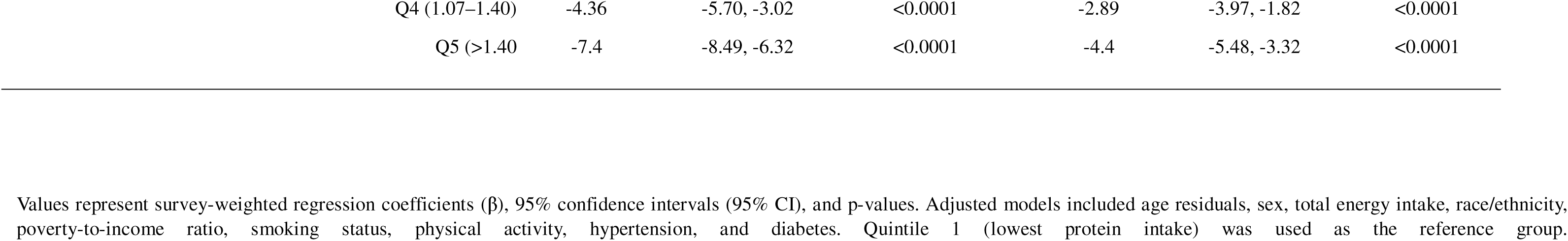
Association between protein intake and biological aging markers.

| Outcome | Exposure | *Unadjusted* |  |  | *Adjusted* |  |  |
| --- | --- | --- | --- | --- | --- | --- | --- |
| Aging Marker | Protein Intake | $\beta$ | 95% CI | *p* | $\beta$ | 95% CI | *p* |
| Telomere length<br>(T/S ratio) | Continuous (g/kg/day) | 0.056 | 0.041, 0.072 | <0.0001 | 0.056 | 0.029, 0.084 | 0.0005 |
|  | Q1 (<0.62) |  | *Reference* |  |  | *Reference* |  |
|  | Q2 (0.62–0.85) | 0.029 | -0.002, 0.060 | 0.067 | 0.053 | 0.013, 0.094 | 0.014 |
|  | Q3 (0.85–1.07) | 0.054 | 0.020, 0.087 | 0.003 | 0.089 | 0.053, 0.125 | 0.0001 |
|  | Q4 (1.07–1.40) | 0.044 | 0.021, 0.067 | 0.0006 | 0.068 | 0.032, 0.104 | 0.001 |
|  | Q5 (>1.40) | 0.082 | 0.057, 0.107 | <0.0001 | 0.093 | 0.055, 0.131 | 0.0001 |
| AnthropoAge<br>(years) | Continuous (g/kg/day) | -6.13 | -6.86, -5.39 | <0.0001 | -4.49 | -5.20, -3.78 | <0.0001 |
|  | Q1 (<0.62) |  | *Reference* |  |  | *Reference* |  |
|  | Q2 (0.62–0.85) | -0.49 | -2.19, 1.22 | 0.561 | -1.76 | -2.28, -1.24 | <0.0001 |
|  | Q3 (0.85–1.07) | -2.36 | -3.81, -0.90 | 0.003 | -3.41 | -3.97, -2.85 | <0.0001 |
|  | Q4 (1.07–1.40) | -3.91 | -6.14, -1.68 | 0.001 | -4.15 | -4.79, -3.50 | <0.0001 |
|  | Q5 (>1.40) | -8.05 | -9.45, -6.65 | <0.0001 | -6.27 | -7.12, -5.41 | <0.0001 |
| PhenoAge<br>(years) | Continuous (g/kg/day) | -6.55 | -7.30, -5.81 | <0.0001 | -5.25 | -5.67, -4.83 | <0.0001 |
|  | Q1 (<0.62) |  | *Reference* |  |  | *Reference* |  |
|  | Q2 (0.62–0.85) | -0.24 | -1.91, 1.44 | 0.773 | -1.67 | -2.35, -0.99 | 0.0001 |
|  | Q3 (0.85–1.07) | -1.91 | -3.69, -0.13 | 0.037 | -2.84 | -3.43, -2.24 | <0.0001 |
|  | Q4 (1.07–1.40) | -3.85 | -6.17, -1.54 | 0.002 | -4.15 | -4.75, -3.56 | <0.0001 |
|  | Q5 (>1.40) | -8.25 | -9.67, -6.83 | <0.0001 | -6.58 | -7.20, -5.96 | <0.0001 |
| KDM<br>(years) | Continuous (g/kg/day) | -5.4 | -6.07, -4.73 | <0.0001 | -3.26 | -4.03, -2.48 | <0.0001 |
|  | Q1 (<0.62) |  | *Reference* |  |  | *Reference* |  |
|  | Q2 (0.62–0.85) | -1.28 | -2.43, -0.13 | 0.03 | -1.23 | -2.10, -0.37 | 0.009 |
|  | Q3 (0.85–1.07) | -2.82 | -4.04, -1.61 | <0.0001 | -2.25 | -3.24, -1.27 | 0.0002 |

|  |  |  |  |  |  |  |
| --- | --- | --- | --- | --- | --- | --- |
| Q4 (1.07–1.40) | -4.36 | -5.70, -3.02 | <0.0001 | -2.89 | -3.97, -1.82 | <0.0001 |
| Q5 (>1.40) | -7.4 | -8.49, -6.32 | <0.0001 | -4.4 | -5.48, -3.32 | <0.0001 |

Values represent survey-weighted regression coefficients ( $\beta$ ), 95% confidence intervals (95% CI), and p-values. Adjusted models included age residuals, sex, total energy intake, race/ethnicity, poverty-to-income ratio, smoking status, physical activity, hypertension, and diabetes. Quintile 1 (lowest protein intake) was used as the reference group.

### Association of dietary protein and BA markers with all-cause and cause-specific mortality

Using Cox models, we found that higher dietary protein intake and longer TL were associated with lower all-cause mortality, whereas higher AnthropoAge, PhenoAge, and KDM were associated with higher mortality. These associations were attenuated after dietary protein intake was included in the models (**Table 3**). For cause-specific mortality, higher protein intake and longer TL were associated with lower cancer mortality, but not heart disease or cerebrovascular mortality (Supplementary Table 8). Likewise, AnthropoAge, PhenoAge, and KDM were associated with heart disease and cancer mortality, with weaker associations for cerebrovascular mortality.

**Table 3.**
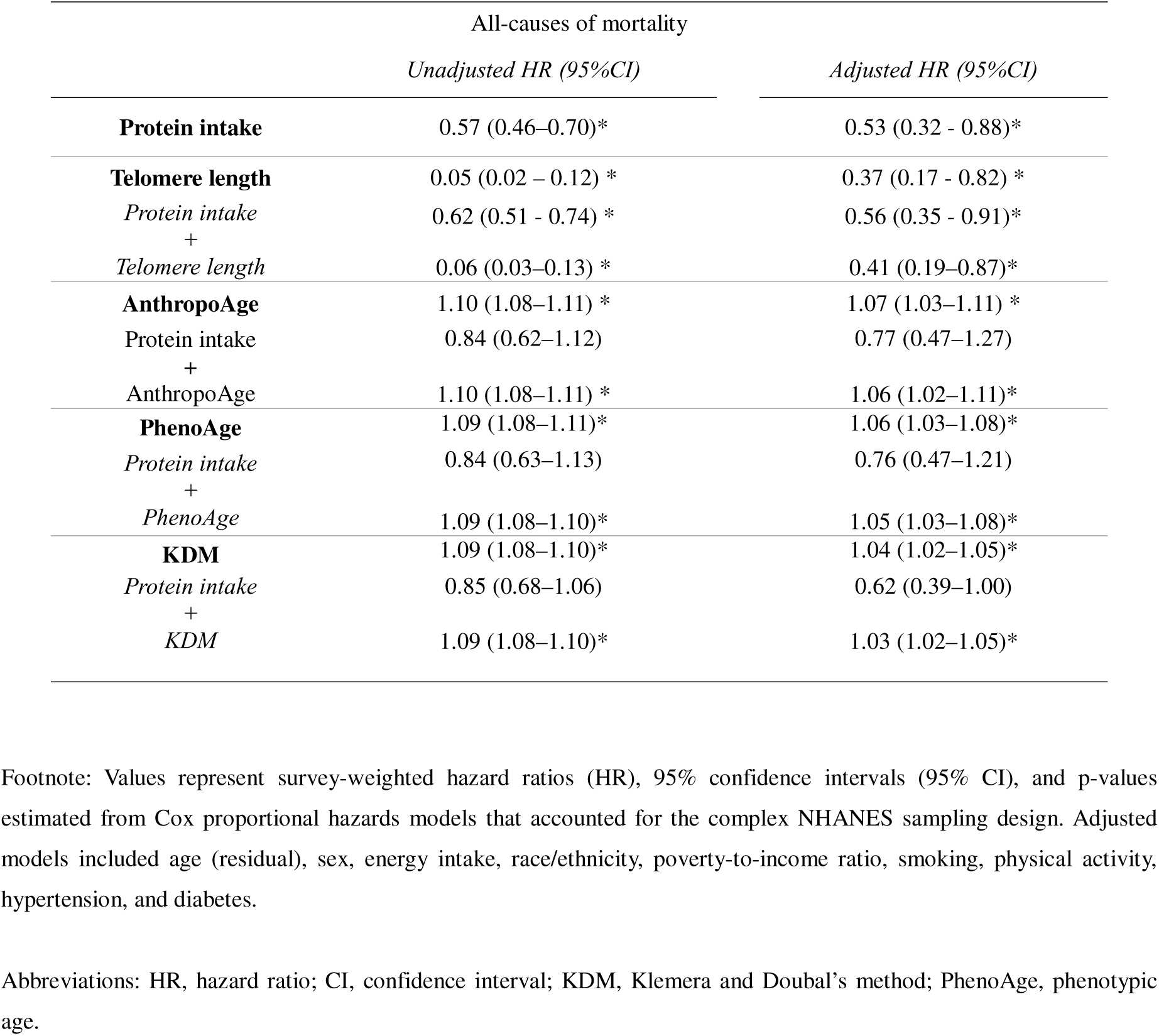
Survey-weighted association between protein intake, biological aging markers, and all-cause mortality.

|  | All-causes of mortality |  |
| --- | --- | --- |
|  | *Unadjusted HR (95%CI)* | *Adjusted HR (95%CI)* |
| **Protein intake** | 0.57 (0.46–0.70)* | 0.53 (0.32 - 0.88)* |
| **Telomere length** | 0.05 (0.02 – 0.12) * | 0.37 (0.17 - 0.82) * |
| Protein intake<br>+ | 0.62 (0.51 - 0.74) * | 0.56 (0.35 - 0.91)* |
| Telomere length | 0.06 (0.03–0.13) * | 0.41 (0.19–0.87)* |
| **AnthropoAge** | 1.10 (1.08–1.11) * | 1.07 (1.03–1.11) * |
| Protein intake<br>+ | 0.84 (0.62–1.12) | 0.77 (0.47–1.27) |
| AnthropoAge | 1.10 (1.08–1.11) * | 1.06 (1.02–1.11)* |
| **PhenoAge** | 1.09 (1.08–1.11)* | 1.06 (1.03–1.08)* |
| Protein intake<br>+ | 0.84 (0.63–1.13) | 0.76 (0.47–1.21) |
| PhenoAge | 1.09 (1.08–1.10)* | 1.05 (1.03–1.08)* |
| **KDM** | 1.09 (1.08–1.10)* | 1.04 (1.02–1.05)* |
| Protein intake<br>+ | 0.85 (0.68–1.06) | 0.62 (0.39–1.00) |
| KDM | 1.09 (1.08–1.10)* | 1.03 (1.02–1.05)* |

Footnote: Values represent survey-weighted hazard ratios (HR), 95% confidence intervals (95% CI), and p-values estimated from Cox proportional hazards models that accounted for the complex NHANES sampling design. Adjusted models included age (residual), sex, energy intake, race/ethnicity, poverty-to-income ratio, smoking, physical activity, hypertension, and diabetes.
Abbreviations: HR, hazard ratio; CI, confidence interval; KDM, Klemmera and Doubal's method; PhenoAge, phenotypic age.

### Mediation analyses

Model-based mediation analyses showed that BA markers partially mediated the association between dietary protein intake and all-cause mortality. TL accounted for 6.9% (95% CI: 6.6–7.3) of the total effect, whereas PhenoAge, AnthropoAge, and KDM accounted for 55.8% (95% CI: 52.0–59.6), 52.7% (95% CI: 47.5–58.2), and 23.1% (95% CI: 22.5–23.7), respectively (**Figure 2**). In cause-specific analyses, mediation was observed primarily for PhenoAge, accounting for 69.1% (95% CI: 59.4–81.5) and 67.1% (95% CI: 57.8–78.8) of the associations between protein intake and heart disease and cancer mortality, respectively. KDM mediated 26.6% (95% CI: 24.3–29.3) of the association between protein intake and heart disease mortality. No clear mediation effects were observed for TL or AnthropoAge, nor for any marker in relation to cerebrovascular mortality (**Supplementary Figure 4**).

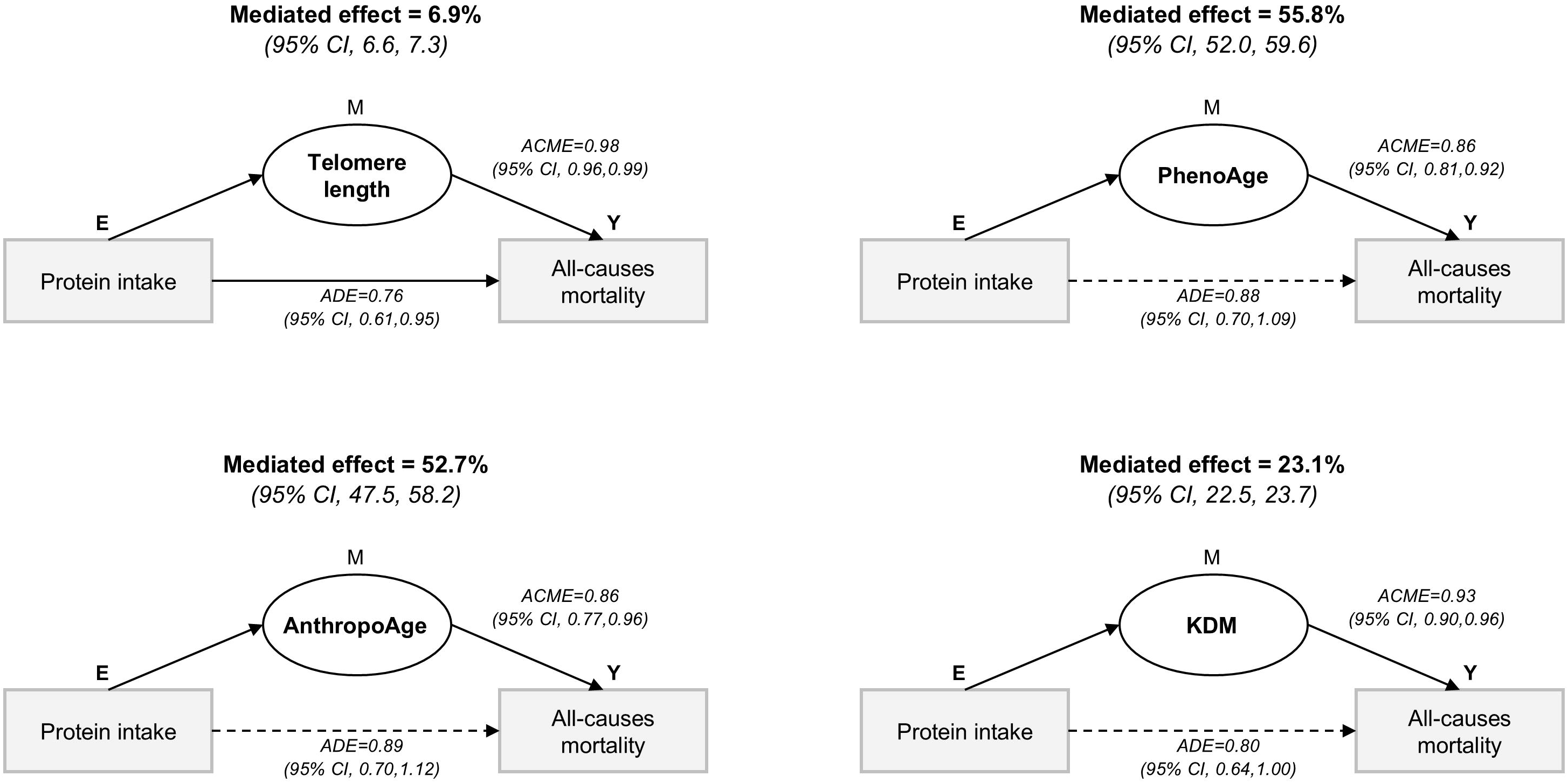

## DISCUSSION

In this study, we evaluated the association between dietary protein intake and BA using TL and three validated BA markers (AnthropoAge, PhenoAge, and KDM) in NHANES 1999–2002 participants. We found that higher dietary protein intake was associated with longer telomeres and lower BA marker values. We also examined their associations with all-cause and cause-specific mortality. Although previous studies have reported an inverse association between protein intake and all-cause mortality, our findings extend this evidence and suggest that BA markers may partially mediate this relationship. Overall, these results support dietary protein intake as a potentially modifiable contributor to healthy aging.

Diet quality is a determinant of health and longevity, as it directly impacts disease and mortality (31). Among dietary components, dietary protein intake plays a crucial role in maintaining tissues, regulating the immune response, and controlling metabolism (32). Evidence supporting its role in aging has grown, particularly because dietary protein intake is a modifiable lifestyle factor that may influence healthy aging (10,32). *Wang et al*. proposed that accelerated BA may serve as a mechanistic link between poor diet quality and chronic diseases (10). In this regard, several validated BA markers have been developed to capture physiological correlations of chronic diseases. Among them, TL is the most widely used molecular marker of aging due to its link to cellular replicative capacity and oxidative stress (33). In our study, we found an association between higher dietary protein intake and TL, particularly when examining a composite of telomere-based age acceleration (TLAgeAccel). Our results align with previous studies that report the protective effects of moderately higher protein intake on TL (34,35). However, TL remains limited in clinical practice due to high variability in measurement and interpretation, as well as the high cost of measurement (6). To overcome these limitations, several composite indices of BA have been proposed (8,20,21). These biomarkers were designed to estimate BA but differ in their underlying components. For example, PhenoAge integrates biochemical markers associated with mortality risk, while AnthropoAge focuses on anthropometric measurements, and KDM estimates aging based on biomarkers associated with CA. These markers may capture distinct aging domains, including body composition, energy regulation, homeostasis, and neurodegeneration/neuroplasticity (36). Our results show an inverse association between higher dietary protein intake and lower BA, particularly for AnthropoAge and PhenoAge, suggesting that dietary protein intake may influence these aging domains.

We found that dietary protein intake was associated with lower all-cause mortality, consistent with previous studies (37,38). Unlike studies focusing on specific protein sources or older populations, our findings extend this evidence by suggesting that BA markers partially mediate this association. Additionally, shorter TL and higher AnthropoAge, PhenoAge, and KDM were associated with increased all-cause mortality, consistent with previous reports (21,23,39). In cause-specific analyses, associations between dietary protein intake and BA markers were observed only for cancer mortality, suggesting that the relationship between protein intake and mortality may vary by cause of death. However, these findings should be interpreted cautiously and require replication, given the limited number of events.

Finally, to explore the mechanisms underlying this relationship, we conducted mediation analyses to help us propose a potential mechanistic pathway. We found that PhenoAge and AnthropoAge explained a substantial proportion of the association between dietary protein intake and all-cause mortality, whereas TL accounted for a relatively small proportion. These findings suggest that while BA may partially explain the observed association between protein intake and mortality, its role in specific causes of death may be more nuanced and pathway dependent. Our results support the view that dietary protein may be associated with lower BA levels across multiple domains, as reflected in both TL and three BA metrics. While our study did not assess differential effects by plant- or animal-based protein sources, prior studies suggest that higher intake of plant-based protein may be more beneficial for BA For example, *Xu et al.* reported that higher plant protein intake was associated with lower mortality (42), whereas *Chen et al.* found associations with lower BA markers that were not observed for animal protein (43). Overall, our findings contribute to the discussion of which protein sources may confer protection against accelerated aging. Future studies should further investigate whether the observed relationships differ by protein quality and origin and confirm their association through intervention designs.

### Strengths and Limitations

Our study provides insights into the association between dietary protein intake and BA. A major strength is the use of a nationally representative survey including TL measurements, which are rarely available in large population-based studies. The inclusion of three BA biomarkers, particularly AnthropoAge, which has not previously been examined in relation to dietary protein intake to our knowledge, and their incorporation into mediation models expands previous evidence and supports a potential mechanistic link between dietary protein intake and mortality. However, some limitations should be acknowledged. First, cross-sectional assessments preclude the precise establishment of temporal relationships, so reverse causality cannot be ruled out. Future longitudinal studies are needed to determine whether increased protein intake precedes and influences BA rather than being a consequence of it. Second, BA markers such as TL, KDM, PhenoAge, and AnthropoAge have been developed and validated mainly in middle-aged and older adults (21,44). As a result, these measures may show limited variability in younger adults, who often have not yet accumulated substantial physiological dysregulation during their lifespan. This limited variability could reduce the ability to detect the associations we explored in younger participants (45). Also, the biological role of dietary protein may differ across the life course. In younger adults, it primarily acts as an anabolic stimulus, while in older adults, it helps protect against sarcopenia and functional decline (46,47). Third, the sample size was limited for mortality outcomes, reducing the statistical power to elucidate specific causes of death. Fourth, dietary protein intake was assessed using a single 24-hour recall, which may not fully capture habitual intake; however, this approach is standard in large-scale population surveys. Fifth, protein intake was normalized to actual body weight, which could underestimate relative protein intake among individuals with overweight or obesity, as current dietary recommendations are based on reference body weights corresponding to normal BMI ranges (48). Future studies should explore whether the use of reference or adjusted body weight provides complementary information regarding the association between protein intake and biological aging. Sixth, the NHANES cycles that we analyzed lacked detailed information about protein sources, preventing us from assessing whether animal- and plant-based proteins contributed differently to the observed effects. However, future studies that specifically differentiate between animal- and plant-based proteins are needed to elucidate how different protein sources could potentially influence cardiometabolic health and aging trajectories. Seventh, the NHANES cycles used for our analyses were conducted over two decades ago (1999–2002). Since then, the U.S. population has undergone demographic and nutritional changes, including an aging population, increased ethnic diversity, and evolving dietary behaviors (49). However, data from NHANES 2005–2018 indicate that total protein intake has remained relatively stable (50). Therefore, despite these contextual shifts, our findings are likely applicable to current dietary patterns in the U.S. Finally, mediation analyses should be interpreted as a statistical decomposition of survival models. Therefore, these results should be interpreted with caution and need to be replicated in prospective studies with a higher number of mortality events.

## CONCLUSION

In this study, using NHANES data, we found that higher dietary protein intake was associated with longer TL and lower BA, as assessed by AnthropoAge, PhenoAge, and KDM. Furthermore, higher dietary protein intake was associated with lower all-cause mortality, with TL and these three BA markers accounting for a proportion of this association. Overall, our findings highlight the interplay between diet, accelerated aging, and mortality risk, as well as the importance of considering potential pathways when evaluating the long-term effects of nutritional exposures. This approach may reinforce dietary recommendations and inform population-level interventions to promote healthy aging.

## Abbreviations

AnthropoAge: Anthropometric Age,
BA: Biological aging,
CA: Chronological age,
KDM: Klemera-Doubal Method,
PhenoAge: Phenotypic Age,
TL: Telomere length.

## ACKNOWLEDGMENTS

The corresponding author confirms the accuracy of authorship, author order, and affiliations, and that all contributors have been appropriately acknowledged. Carlos A. Fermín-Martínez is enrolled in the PECEM Program at the Faculty of Medicine, UNAM, and is supported by CONACyT.

## AUTHOR CONTRIBUTIONS

JRB and NEAV conceived this study; JRB, NEAV, and OYBC harmonized the dataset; JRB, NEAV, and OYBC conducted the analysis; JRB, NEAV, CAFM, AKG, NASO, and RRR drafted the first version of the manuscript; NEAV, OYBC, CAFM, AKG, NASO, RRR provided mentorship and guidance. Each author contributed important intellectual content during manuscript drafting or revision and accepted accountability for the overall work by ensuring that questions about the accuracy or integrity of any portion of the work were appropriately investigated and resolved.

## DATA AVAILABILITY

All code, datasets, and materials necessary for the reproducibility of results are available at: NHANES datasets: Publicly available NHANES datasets can be downloaded from: https://wwwn.cdc.gov/nchs/nhanes/continuousnhanes/default.aspx. Mortality data: Linked mortality data from NHANES can be accessed at: https://www.cdc.gov/nchs/data-linkage/mortality-public.html

## ETHICAL STATEMENT

The authors declare that all experiments involving human subjects were conducted in accordance with the Declaration of Helsinki (http://www.wma.net) and that all procedures were carried out with the subjects’ adequate understanding and written consent.

