## Supplementary Material for "Association of dietary protein intake with accelerated biological aging markers and all-cause mortality in adults"

**INDEX**

Supplementary Figure 1*: Directed acyclic graphs for the relationship between protein intake and biological aging markers.*

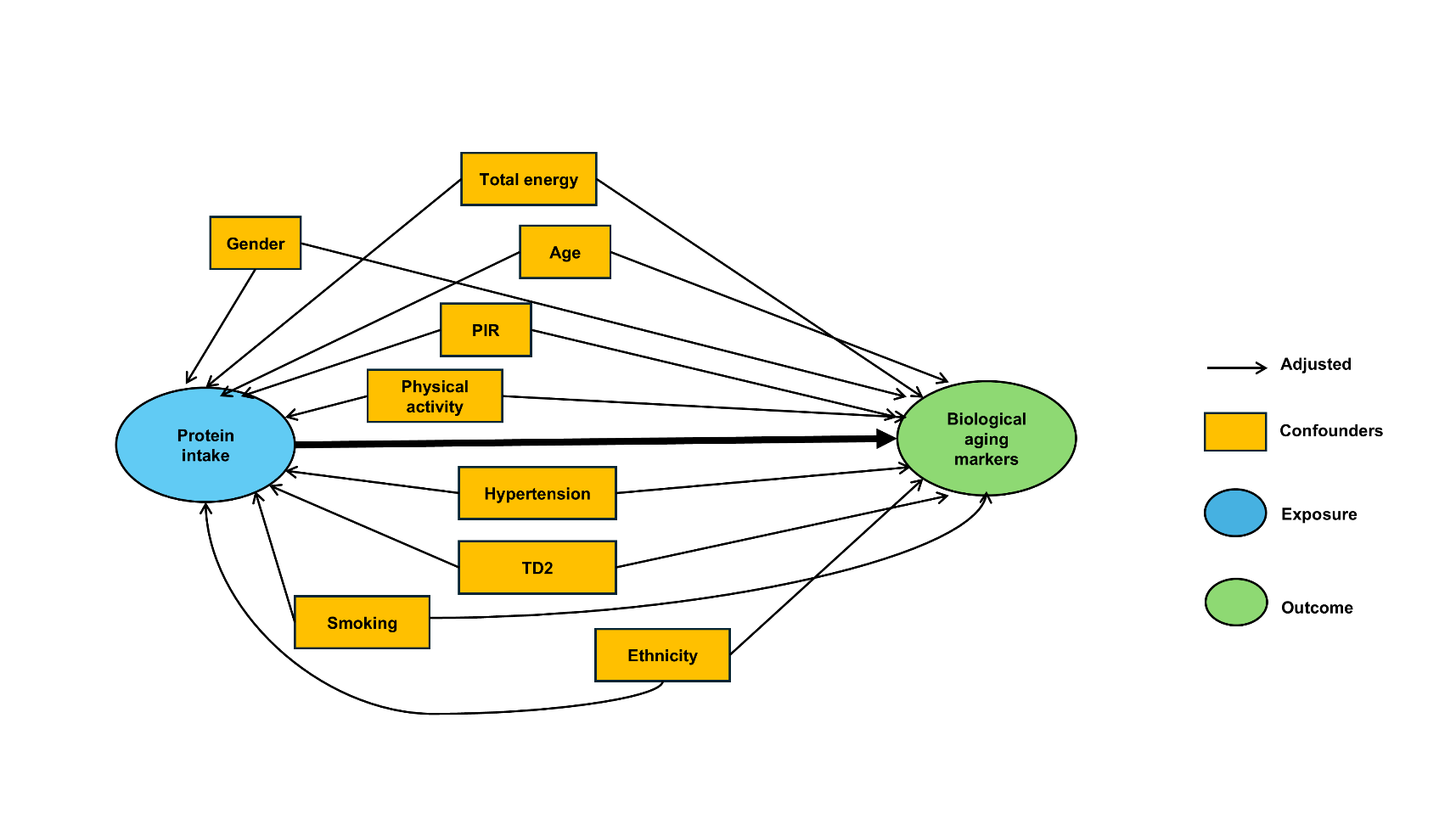

*Abbreviations: PIR, poverty-to-income ratio; T2D, type 2 diabetes.*

Supplementary Figure 2***:*** *STROBE flowchart displaying the sample selection from NHANES.*

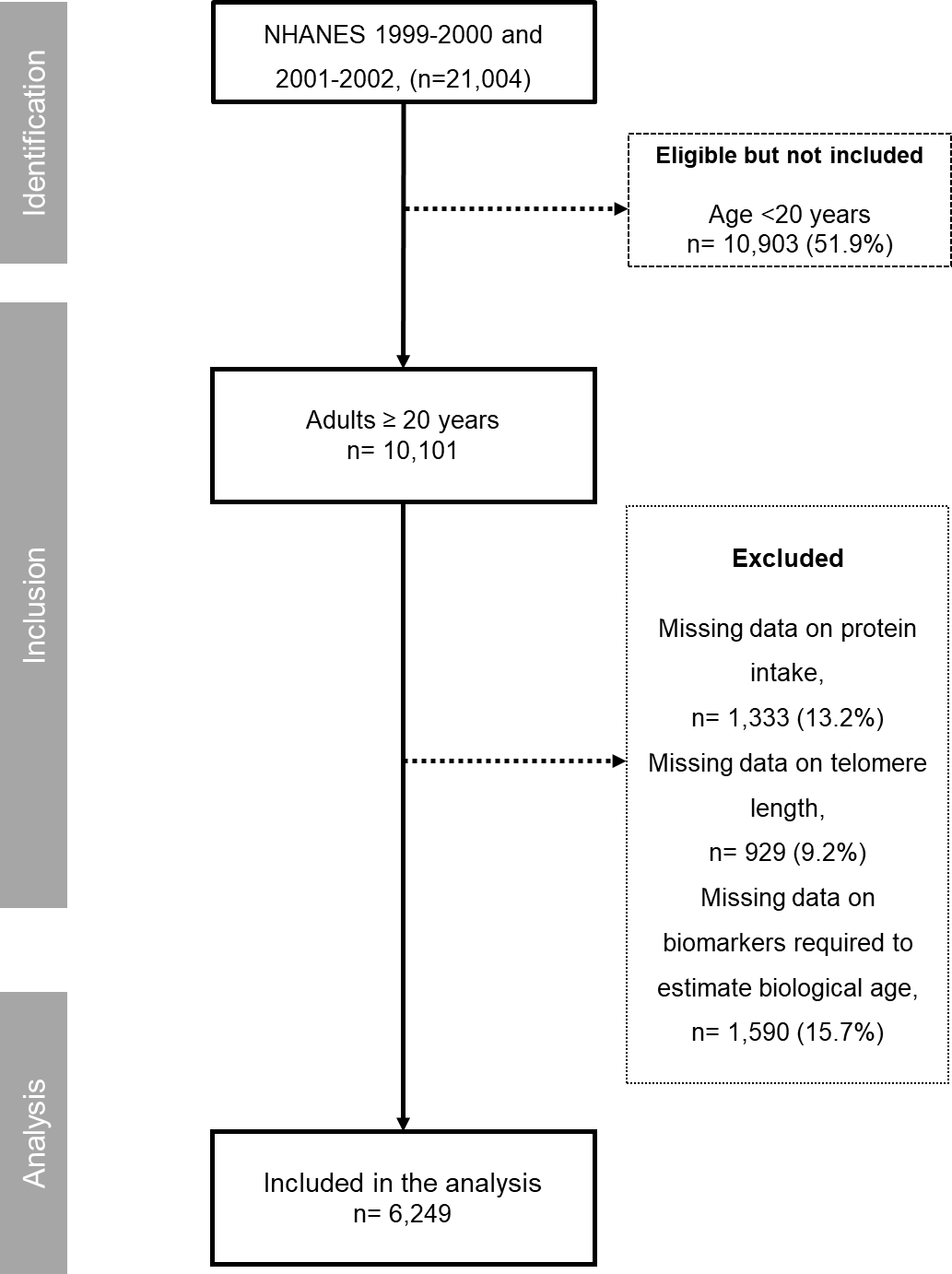

*Abbreviations: NHANES, National Health and Nutrition Examination Survey*

Supplementary Figure 3*:* *Weighted correlation matrix and scatterplots showing the association between protein intake and biological aging markers.*

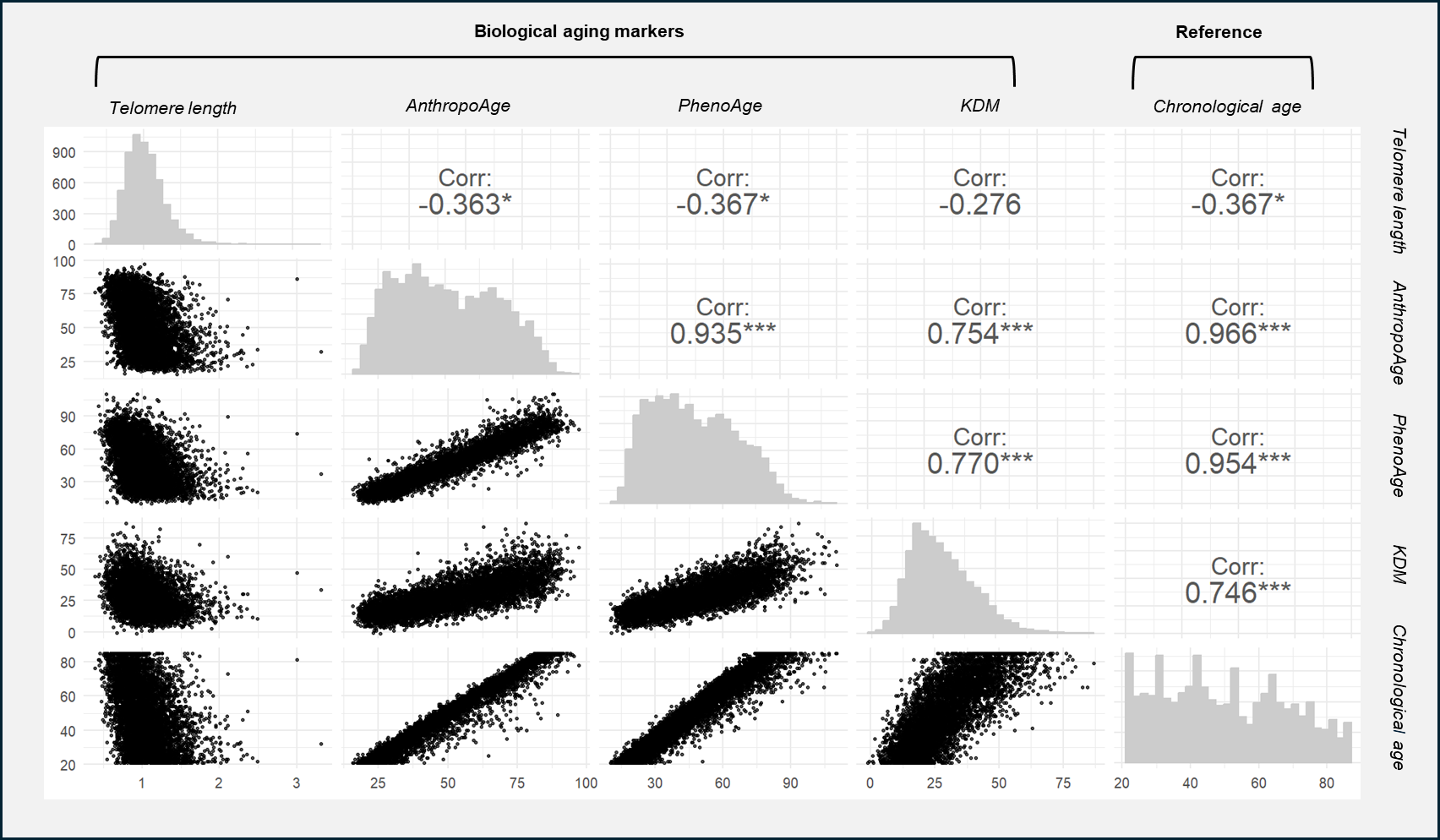

*Correlation coefficients were estimated using weighted Pearson correlations accounting for the complex survey design.
* p < 0.05, **** ***p*** *< 0.001.*

Abbreviations: AnthropoAge, anthropometric age; PhenoAge, phenotypic age; KDM, Klemera and Doubal’s method.

Supplementary Figure 4*: Mediation analysis of the association between protein intake and overall and cause-specific mortality through aging markers*

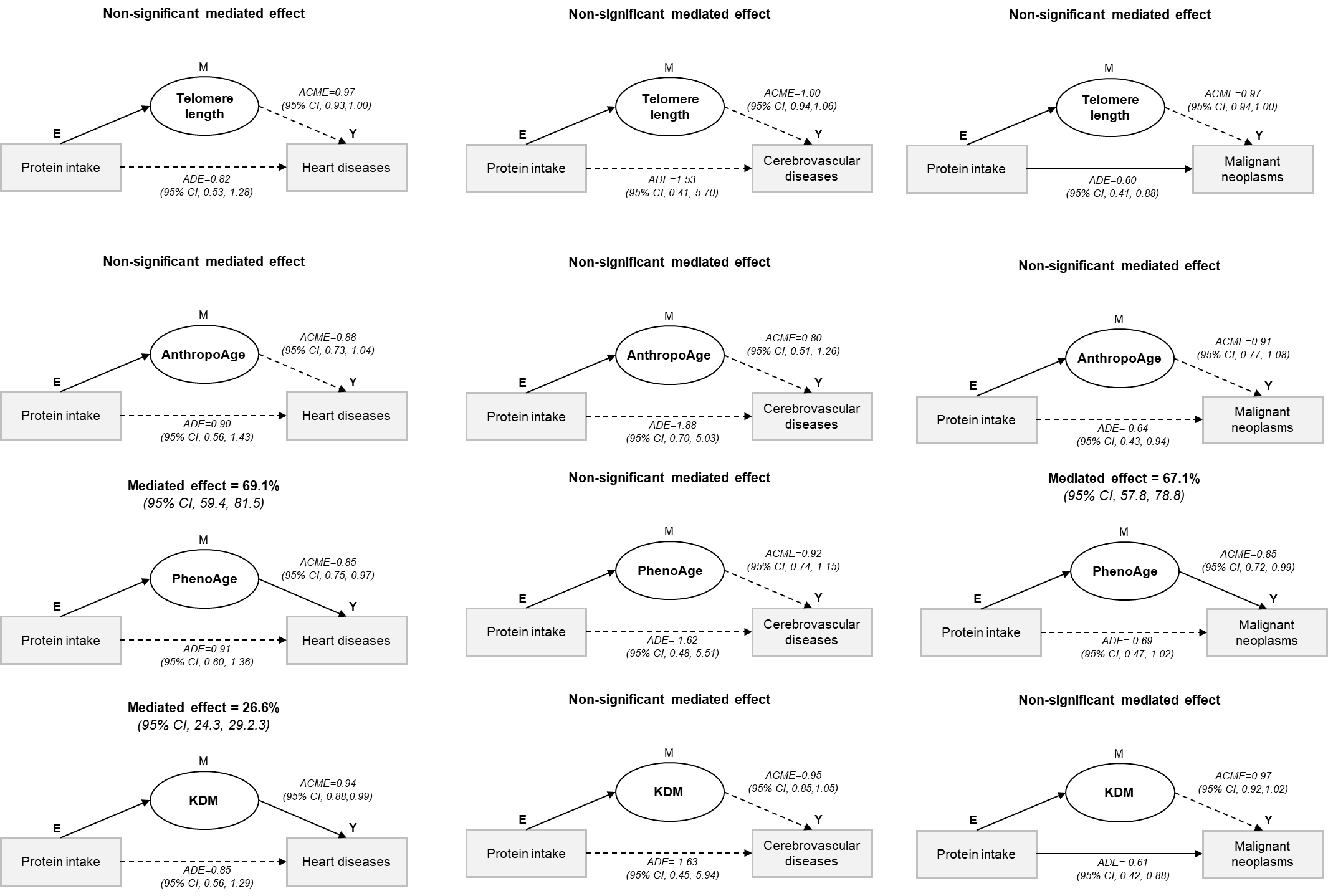

*Footnote: ACME (average causal mediation effect) represents the indirect effect, and ADE (average direct effect) represents the direct effect. Values are hazard ratios (HR) with 95% confidence intervals (CI). Mediation analyses were conducted using survey-weighted regression models accounting for the complex NHANES sampling design (strata, clusters, and examination weights). The proportion mediated was calculated on the log-hazard scale as log(ACME)/log(total effect), with 95% confidence intervals derived using the delta method. Models were adjusted for age (residual), sex, energy intake (kcal), race/ethnicity, poverty-to-income ratio, smoking, physical activity, hypertension, and diabetes.*).

Supplementary Table 1*: STROBE reporting checklist for the study.*

|  | Item Description | Location (or reason for not reporting) |
| --- | --- | --- |
| **Title and abstract** |  |  |
| [1a. Indicate the study’s design](https:/resources.equator-network.org/reporting-guidelines/strobe/items/title-abstract-indicate-study-design.html?utm_source=strobe&utm_medium=checklist&utm_campaign=1_1) | Indicate the study’s design with a commonly used term in the title or the abstract. | Title page |
| [1b. Abstract](https:/resources.equator-network.org/reporting-guidelines/strobe/items/abstract.html?utm_source=strobe&utm_medium=checklist&utm_campaign=1_1) | Provide in the abstract an informative and balanced summary of what was done and what was found. | Page 1 |
| **Introduction** |  |  |
| [2. Background / rationale](https:/resources.equator-network.org/reporting-guidelines/strobe/items/background-rationale.html?utm_source=strobe&utm_medium=checklist&utm_campaign=1_1) | Explain the scientific background and rationale for the investigation being reported. | Page 3 |
| [3. Objectives](https:/resources.equator-network.org/reporting-guidelines/strobe/items/objectives.html?utm_source=strobe&utm_medium=checklist&utm_campaign=1_1) | State specific objectives, including any prespecified hypotheses. | Page 3 |
| **Methods** |  |  |
| [4. Study design](https:/resources.equator-network.org/reporting-guidelines/strobe/items/study-design.html?utm_source=strobe&utm_medium=checklist&utm_campaign=1_1) | Present key elements of study design early in the paper. | Pages 6 -9 |
| [5. Setting](https:/resources.equator-network.org/reporting-guidelines/strobe/items/setting.html?utm_source=strobe&utm_medium=checklist&utm_campaign=1_1) | Describe the setting, locations, and relevant dates, including periods of recruitment, exposure, follow-up, and data collection. | Pages 6 -9 |
| [6a. Eligibility criteria](https:/resources.equator-network.org/reporting-guidelines/strobe/items/eligibility-criteria.html?utm_source=strobe&utm_medium=checklist&utm_campaign=1_1) | **Cohort study:** Give the eligibility criteria, and the sources and methods of selection of participants. Describe methods of follow-up. **Case-control study:** Give the eligibility criteria, and the sources and methods of case ascertainment and control selection. Give the rationale for the choice of cases and controls. **Cross-sectional study:** Give the eligibility criteria, and the sources and methods of selection of participants. | Pages 6 -9 |
| [6b. Matching criteria](https:/resources.equator-network.org/reporting-guidelines/strobe/items/matching-criteria.html?utm_source=strobe&utm_medium=checklist&utm_campaign=1_1) | **Cohort study:** For matched studies, give matching criteria and number of exposed and unexposed. **Case-control study:** For matched studies, give matching criteria and the number of controls per case. | Pages 6 -9 |
| [7. Variables](https:/resources.equator-network.org/reporting-guidelines/strobe/items/variables.html?utm_source=strobe&utm_medium=checklist&utm_campaign=1_1) | Clearly define all outcomes, exposures, predictors, potential confounders, and effect modifiers. Give diagnostic criteria, if applicable. | Pages 6 -9 |
| [8. Data sources / measurement](https:/resources.equator-network.org/reporting-guidelines/strobe/items/data-sources-measurement.html?utm_source=strobe&utm_medium=checklist&utm_campaign=1_1) | For each variable of interest give sources of data and details of methods of assessment (measurement). Describe comparability of assessment methods if there is more than one group. | Pages 6 -9 |
| [9. Bias](https:/resources.equator-network.org/reporting-guidelines/strobe/items/bias.html?utm_source=strobe&utm_medium=checklist&utm_campaign=1_1) | Describe any efforts to address potential sources of bias. | Pages 6 -9 |
| [10. Study size](https:/resources.equator-network.org/reporting-guidelines/strobe/items/study-size.html?utm_source=strobe&utm_medium=checklist&utm_campaign=1_1) | Explain how the study size was arrived at. | Pages 6 -9 |
| [11. Quantitative variables](https:/resources.equator-network.org/reporting-guidelines/strobe/items/quantitative-variables.html?utm_source=strobe&utm_medium=checklist&utm_campaign=1_1) | Explain how quantitative variables were handled in the analyses. If applicable, describe which groupings were chosen, and why. | Pages 6 -9 |
| [12a. Statistical methods](https:/resources.equator-network.org/reporting-guidelines/strobe/items/statistical-methods-description.html?utm_source=strobe&utm_medium=checklist&utm_campaign=1_1) | Describe all statistical methods, including those used to control for confounding. | Pages 10-12 |
| [12b. Statistical methods – subgroups and interactions](https:/resources.equator-network.org/reporting-guidelines/strobe/items/statistical-methods-subgroups-interactions.html?utm_source=strobe&utm_medium=checklist&utm_campaign=1_1) | Describe any methods used to examine subgroups and interactions. | Pages 10-12 |
| [12c. Statistical methods – missing data](https:/resources.equator-network.org/reporting-guidelines/strobe/items/statistical-methods-missing-data.html?utm_source=strobe&utm_medium=checklist&utm_campaign=1_1) | Explain how missing data were addressed. | Pages 10-12 |
| [12di. Statistical methods – loss to follow-up](https:/resources.equator-network.org/reporting-guidelines/strobe/items/statistical-methods-loss-to-follow-up.html?utm_source=strobe&utm_medium=checklist&utm_campaign=1_1) | **Cohort study:** If applicable, describe how loss to follow-up was addressed. | Pages 10-12 |
| [12dii. Statistical methods – matching cases and controls](https:/resources.equator-network.org/reporting-guidelines/strobe/items/statistical-methods-matching-cases-controls.html?utm_source=strobe&utm_medium=checklist&utm_campaign=1_1) | **Case-control study:** If applicable, explain how matching of cases and controls was addressed. | NA |
| [12diii. Statistical methods – sampling strategy](https:/resources.equator-network.org/reporting-guidelines/strobe/items/statistical-methods-analytical-methods-sampling-strategy.html?utm_source=strobe&utm_medium=checklist&utm_campaign=1_1) | **Cross-sectional study:** If applicable, describe analytical methods taking account of sampling strategy. | Pages 10-12 |
| [12e. Statistical methods – sensitivity analyses](https:/resources.equator-network.org/reporting-guidelines/strobe/items/statistical-methods-sensitivity-analyses.html?utm_source=strobe&utm_medium=checklist&utm_campaign=1_1) | Describe any sensitivity analyses. | Pages 10-12 |
| **Results** |  |  |
| [13a. Participant numbers](https:/resources.equator-network.org/reporting-guidelines/strobe/items/participants-numbers.html?utm_source=strobe&utm_medium=checklist&utm_campaign=1_1) | Report the numbers of individuals at each stage of the study—e.g., numbers potentially eligible, examined for eligibility, confirmed eligible, included in the study, completing follow-up, and analysed; Consider use of a flow diagram. | Pages 12-16 |
| [13b. Participants – non-participation](https:/resources.equator-network.org/reporting-guidelines/strobe/items/participants-non-participation.html?utm_source=strobe&utm_medium=checklist&utm_campaign=1_1) | Give reasons for non-participation at each stage. | Pages 12-16 |
| [13c. Participants – flow diagram](https:/resources.equator-network.org/reporting-guidelines/strobe/items/participants-flow-diagram.html?utm_source=strobe&utm_medium=checklist&utm_campaign=1_1) | Consider use of a flow diagram. | Figure 1 |
| [14a. Descriptive data – participant characteristics](https:/resources.equator-network.org/reporting-guidelines/strobe/items/descriptive-data-participant-characteristics.html?utm_source=strobe&utm_medium=checklist&utm_campaign=1_1) | Give characteristics of study participants (e.g., demographic, clinical, social) and information on exposures and potential confounders. Present the information in a table. | Pages 12-16 |
| [14b. Descriptive data – missing data](https:/resources.equator-network.org/reporting-guidelines/strobe/items/descriptive-data-missing-data.html?utm_source=strobe&utm_medium=checklist&utm_campaign=1_1) | Indicate the number of participants with missing data for each variable of interest. | Pages 12-16 |
| [14c. Descriptive data – follow-up time](https:/resources.equator-network.org/reporting-guidelines/strobe/items/descriptive-data-follow-up-time.html?utm_source=strobe&utm_medium=checklist&utm_campaign=1_1) | **Cohort study:** Summarise follow-up time—e.g., average and total amount. | Pages 12-16 |
| [15. Outcome data](https:/resources.equator-network.org/reporting-guidelines/strobe/items/outcome-data.html?utm_source=strobe&utm_medium=checklist&utm_campaign=1_1) | **Cohort study:** Report numbers of outcome events or summary measures over time. **Case-control study:** Report numbers in each exposure category, or summary measures of exposure. **Cross-sectional study:** Report numbers of outcome events or summary measures. | Pages 12-16 |
| [16a. Main results](https:/resources.equator-network.org/reporting-guidelines/strobe/items/main-results.html?utm_source=strobe&utm_medium=checklist&utm_campaign=1_1) | Give unadjusted estimates and, if applicable, confounder-adjusted estimates and their precision (e.g., 95% confidence intervals). Make clear which confounders were adjusted for and why they were included. | Pages 12-16 |
| [16b. Main results – category boundaries](https:/resources.equator-network.org/reporting-guidelines/strobe/items/main-results-category-boundaries.html?utm_source=strobe&utm_medium=checklist&utm_campaign=1_1) | Report category boundaries when continuous variables were categorised. | Pages 12-16 |
| [16c. Main results – risk](https:/resources.equator-network.org/reporting-guidelines/strobe/items/main-results-risk.html?utm_source=strobe&utm_medium=checklist&utm_campaign=1_1) | If relevant, consider translating estimates of relative risk into absolute risk for a meaningful time period. | Pages 12-16 |
| [17. Other analyses](https:/resources.equator-network.org/reporting-guidelines/strobe/items/other-analyses.html?utm_source=strobe&utm_medium=checklist&utm_campaign=1_1) | Report other analyses done—e.g., analyses of subgroups and interactions, and sensitivity analyses. | Pages 12-16 |
| **Discussion** |  |  |
| [18. Key results](https:/resources.equator-network.org/reporting-guidelines/strobe/items/key-results.html?utm_source=strobe&utm_medium=checklist&utm_campaign=1_1) | Summarise key results with reference to study objectives. | Pages 17-22 |
| [19. Limitations](https:/resources.equator-network.org/reporting-guidelines/strobe/items/limitations.html?utm_source=strobe&utm_medium=checklist&utm_campaign=1_1) | Discuss limitations of the study, taking into account sources of potential bias or imprecision. Discuss both direction and magnitude of any potential bias. | Pages 17-22 |
| [20. Interpretation](https:/resources.equator-network.org/reporting-guidelines/strobe/items/interpretation.html?utm_source=strobe&utm_medium=checklist&utm_campaign=1_1) | Give a cautious overall interpretation considering objectives, limitations, multiplicity of analyses, results from similar studies, and other relevant evidence. | Pages 17-22 |
| [21. Generalisability](https:/resources.equator-network.org/reporting-guidelines/strobe/items/generalisability.html?utm_source=strobe&utm_medium=checklist&utm_campaign=1_1) | Discuss the generalisability (external validity) of the study results. | Pages 17-22 |
| **Other information** |  |  |
| [22. Funding](https:/resources.equator-network.org/reporting-guidelines/strobe/items/funding.html?utm_source=strobe&utm_medium=checklist&utm_campaign=1_1) | Give the source of funding and the role of the funders for the present study and, if applicable, for the original study on which the present article is based. | Page 28 |

*Abbreviations: STROBE, Strengthening the Reporting of Observational Studies in Epidemiology.*

Supplementary Table 2*: Methodology for dietary intake assessment in NHANES 1999–2000 and 2001–2002.*

| **Aspect** | **Description** |
| --- | --- |
| Study Description | *NHANES comprises a series of cross-sectional surveys conducted in the United States since the 1960s by the National Center for Health Statistics (NCHS), a component of the Centers for Disease Control and Prevention (CDC).* |
| Ethics Approval | *NHANES has received approval from the NCHS Ethics Committee.* |
| Survey | National Health and Nutrition Examination Survey (NHANES) 1999–2000 and 2001-2002. |
| Data Files Used | Total Nutrient Intakes File (DRXTOT) |
| Objective | Obtain detailed dietary intake information to estimate total intake of energy, macronutrients, and non-nutrient food components from foods and beverages consumed in the 24-hour period prior to the interview. |
| Dietary Assessment Method | 24-hour dietary recall interview (midnight-to-midnight), using the USDA Automated Multiple-Pass Method (AMPM), computer-assisted (CADI system). |
| Interview Mode | Primarily in-person at Mobile Examination Centers (MEC); a small subset conducted by telephone (Dietary Interview Mode Evaluation Study). |
| Respondent Eligibility | All examinees; proxy interviews for children <6 years; assisted interviews for ages 6–11 years. |
| Survey Staff | Trained bilingual interviewers with B.S. in Nutrition/Home Economics; 2-week training plus supervised practice. |
| Data Collection Components | • Quick List: recall all foods and beverages consumed. • Time, Occasion, Place: record time, location, and meal type. • Food Details: probes for preparation, brand, portion size. • Final Review: review and confirm all reported foods. • Post-Recall Questionnaire: water intake, fish/shellfish frequency. |
| Coding & Data Processing | Imported into University of Texas Food Intake Analysis System (FIAS v3.99) with USDA 1994–98 nutrient database. Modifications for recipe changes, foods not in database, and sodium adjustments applied. |
| Quality Control | • Audio review of ~5% of interviews. • Review for missing data or inconsistencies. • Double coding of 10% of foods. • Staff retraining as needed. |
| Exclusions | Implausible intake (<500 kcal/day or >5,000 kcal/day), missing dietary data, infants/children with unquantified human milk intake. |
| Weighting | Sample weights (WTDRD1 for 2-year dietary data) are used to produce nationally representative estimates. Adjustments for non-response and day-of-week differences applied. |
| References | (1) |

Abbreviations: NHANES, National Health and Nutrition Examination Survey; NCHS, National Center for Health Statistics; CDC, Centers for Disease Control and Prevention; MEC, Mobile Examination Center; AMPM, Automated Multiple-Pass Method; CADI, Computer-Assisted Dietary Interview; FIAS, Food Intake Analysis System; USDA, United States Department of Agriculture; DRXTOT, Total Nutrient Intakes File.

Supplementary Table 3*:* Methodology for telomere length determination in NHANES 1999–2000 and 2001–2002.

| **Aspect** | **Description** |
| --- | --- |
| Study Description | *NHANES comprises a series of cross-sectional surveys conducted in the United States since the 1960s by the National Center for Health Statistics (NCHS), a component of the Centers for Disease Control and Prevention (CDC).* |
| Ethics Approval | *NHANES has received approval from the NCHS Ethics Committee.* |
| Survey | National Health and Nutrition Examination Survey (NHANES) 1999–2000 (and 2001–2002 for extended analysis). |
| Data File Used | TELO_A.xpt (Telomere Mean and Standard Deviation, Surplus data file). |
| Eligible Sample | Participants aged ≥20 years with available blood samples for DNA purification. |
| Sample Collection | Blood collected at Mobile Examination Centers (MEC); DNA extracted and stored for later analysis. |
| Laboratory | Laboratory of Dr. Elizabeth Blackburn, University of California, San Francisco. |
| Assay Method | Quantitative PCR (qPCR) to measure relative telomere length (T/S ratio), as described by Cawthon (2002). |
| Assay Procedure | • Each sample assayed 3 times on 3 different days. • Duplicate wells per assay (6 data points per sample). • Samples run in groups of 3 plates; no plates grouped together more than once. • Each plate included 96 control wells with 8 control DNA samples. |
| Quality Control | • Assay runs with ≥8 invalid control wells excluded (<1%). • Runs with >4 control DNA values outside 2.5 SD from mean excluded (<6%). • Outliers within samples excluded (<2%). • Five blinded QC plates (~5% of total set); ≥95% duplicate concordance required. • Inter-assay coefficient of variation: 6.5%. |
| Data Processing | Mean and standard deviation (SD) of T/S ratio calculated per sample. |
| Analytic Notes | • TeloMean: Mean T/S ratio (relative telomere length). • TeloSTD: Standard deviation of 3 T/S ratio values. • Conversion to base pairs: bp = 3,274 + 2,413 × (T/S). Valid only for assays conducted in the Blackburn Lab under same conditions. |
| Weighting | MEC exam weights appropriate for telomere subsample analyses. |
| References | (2,3) |

Abbreviations: NHANES, National Health and Nutrition Examination Survey; NCHS, National Center for Health Statistics; CDC, Centers for Disease Control and Prevention; MEC, Mobile Examination Center; qPCR, quantitative polymerase chain reaction; T/S ratio, telomere to single-copy gene ratio; SD, standard deviation; QC, quality control; bp, base pairs.

Supplementary table 4: Biological aging markers and their estimation methods.

| **Clinical aging markers** | **Variables used** | **Estimation method** | **Interpretation** | **Reference** |
| --- | --- | --- | --- | --- |
| *AnthropoAge (simplified version)* | Chronological age (CA), Body mass index (BMI), Waist-to-Hip ratio(WHR). | Derived from proportional hazards regression models with the Gompertz distribution. Mortality risk predicted from CA and anthropometry was equated and expressed in years. (AnthropoAge R package) | Estimates biological age from mortality risk associated with anthropometric features reflect physical/structural aging. | (4) |
| *PhenoAge* | Albumin, Alkaline phosphatase, Creatinine, C-reactive protein (CRP), Glucose, Mean cell volume (MCV), Red cell distribution width (RDW), White blood cell count (WBC), Lymphocyte proportion + CA. | Calculated from a linear combination of CA and nine multi-system clinical chemistry biomarkers, optimized to predict all-cause mortality risk (BioAge R package). | Captures biological age from systemic biochemical and hematological markers; reflects physiological aging and health status, strongly associated with morbidity and mortality. | (5) |
| *KDM* | Forced expiratory volume in 1 second (FEV1), Systolic blood pressure, Albumin, Alkaline phosphatase, Blood urea nitrogen (BUN), Creatinine, CRP, HbA1c, Total cholesterol. | Reverse regression method using CA as a reference biomarker. Combines biomarker-specific regressions to compute BA, improving precision by minimizing error variance. Implemented with the *BioAge* R package | Provides an integrated estimate of biological age which mean physiological function matches that of the individual. Designed to enhance accuracy of BA estimation | (6) |

Abbreviations: BA, biological age; CA, chronological age; BMI, body mass index; WHR, waist-to-hip ratio; CRP, C-reactive protein; MCV, mean cell volume; RDW, red cell distribution width; WBC, white blood cell count; BUN, blood urea nitrogen; HbA1c, glycated hemoglobin; AnthropoAge, anthropometric age; PhenoAge, phenotypic age; KDM, Klemera and Doubal’s method.

Supplementary Table 5 Survey-weighted dietary intake of the study population (N = 6,249).

| Variable | Value |
| --- | --- |
| Total energy intake (kcal/day) | 2,048 (1,517–2,725) |
| Energy from carbohydrates (kcal/day) | 1003 (723–1,350) |
| Carbohydrates (% of total kcal) | 50 (43–58) |
| Carbohydrate intake (g/day) | 251 (181–337) |
| Energy from fat (kcal/day) | 665 (456–960) |
| Fat (% of total kcal) | 34 (28–39) |
| Total fat intake (g/day) | 74 (51–107) |
| Energy from protein (kcal/day) | 301 (214–407) |
| Protein (% of total kcal) | 15 (12–18.0) |
| Protein intake (g/day) | 75 (54–102) |
| Protein per kg of body weight (g/kg/day) | 0.95 (0.68–1.30) |
| Protein per kg of ideal body weight (g/kg IBW/day) | 1.09 (0.80–1.45) |
| Protein adequacy | 3,853 (62%) |

Values are presented as survey-weighted medians (interquartile range) for continuous variables and as weighted proportions (%) with unweighted sample sizes (n) for categorical variables. All estimates account for the complex sampling design of NHANES using sample weights, strata, and primary sampling units.

Protein adequacy was defined by a threshold of ≥0.8 g/kg/day. Protein per kg of ideal body weight (g/kg IBW/day) was calculated using an adjusted approach: for individuals with BMI ≥25 kg/m², IBW was estimated as 24.9 × height² (m); for those with BMI <18.5 kg/m², IBW was estimated as 18.5 × height²; and for individuals with normal BMI (18.5–24.9 kg/m²), actual body weight was used as IBW.

Abbreviations: IBW, ideal body weight; BMI, body mass index; IQR, interquartile range; kcal, kilocalories*.*

|  | <40  β (95% CI) | 40–60  β (95% CI) | >60  β (95% CI) | *p*-interaction (40–60 vs <40) | *p*-interaction (>60 vs <40) |
| --- | --- | --- | --- | --- | --- |
| Telomere length (T/S ratio) | 0.041 (-0.022, 0.104) | 0.066 (0.012, 0.121)* | 0.119 (0.057, 0.181)* | 0.54 | 0.09 |
| AnthropoAge (years) | -4.530 (-5.990, -3.070)* | -5.789 (-7.466, -4.112)* | -4.852 (-6.801, -2.903)* | 0.24 | 0.59 |
| PhenoAge (years) | -4.920 (-5.768, -4.071)* | -7.461 (-8.591, -6.331)* | -6.000 (-8.029, -3.972)* | 0.12 | 0.16 |
| KDM (years) | -3.805 (-5.258, -2.353)* | -5.259 (-6.959, -3.559)* | -4.739 (-7.814, -1.665)* | 0.84 | 0.8 |

Supplementary Table 6: Survey-weighted age-stratified associations between protein intake and biological aging markers

Footnote: Associations between protein intake (g/kg/day) and biological aging markers were examined within age strata (<40, 40–60, and >60 years) using survey-weighted linear regression models that accounted for the complex NHANES sampling design. Values represent regression coefficients (β) and 95% confidence intervals (CI). Models were adjusted for sex, residual age, energy intake, race/ethnicity, diabetes, physical activity, hypertension, smoking status, and family poverty-income ratio. Asterisks (*) indicate statistically significant associations (p < 0.05). Interaction p-values were obtained from survey-weighted models including cross-product terms between protein intake and age group, using individuals aged <40 years as the reference group.

Abbreviations: PhenoAge, phenotypic age; AnthropoAge, anthropometric age; T/S ratio, telomere to single-copy gene ratio; β, regression coefficient; CI, confidence interval.

Supplementary Table 7: Survey-weighted association between protein intake and biological aging markers (linear regression models)

| **Outcome** | **Exposure** |  | ***Model 1 ^a^*** | | | ***Model 2 ^b^*** | | |
| --- | --- | --- | --- | --- | --- | --- | --- | --- |
| Aging Marker | Protein Intake |  | **β** | **95% CI** | ***p*** | **β** | **95% CI** | ***p*** |
| Telomere length  (T/S ratio) | *Continuous (g/kg/day)* |  | 0.041 | 0.015, 0.067 | 0.007 | 0.057 | 0.025, 0.090 | 0.003 |
|  | *Q5 (>1.40)* |  | 0.084 | 0.053, 0.114 | <0.0001 | 0.103 | 0.067, 0.139 | <0.0001 |
|  | *Q4 (1.07-1.40)* |  | 0.070 | 0.038, 0.101 | 0.001 | 0.081 | 0.053, 0.109 | <0.0001 |
|  | *Q3 (0.85-1.07)* |  | 0.111 | 0.064, 0.158 | <0.0001 | 0.120 | 0.070, 0.169 | <0.0001 |
|  | *Q2 (0.62-0.85)* |  | 0.081 | 0.027, 0.136 | 0.011 | 0.086 | 0.033, 0.140 | 0.006 |
| AnthropoAge  (years) | *Continuous (g/kg/day)* |  | -5.507 | -6.236, -4.778 | <0.0001 | -4.901 | -5.867, -3.935 | <0.0001 |
|  | *Q5 (>1.40)* |  | -7.812 | -8.798, -6.827 | <0.0001 | -6.796 | -8.062, -5.530 | <0.0001 |
|  | *Q4 (1.07-1.40)* |  | -5.271 | -6.011, -4.531 | <0.0001 | -4.640 | -5.577, -3.703 | <0.0001 |
|  | *Q3 (0.85-1.07)* |  | -4.264 | -4.803, -3.726 | <0.0001 | -3.777 | -4.454, -3.100 | <0.0001 |
|  | *Q2 (0.62-0.85)* |  | -2.385 | -3.004, -1.765 | <0.0001 | -2.081 | -2.716, -1.447 | <0.0001 |
| PhenoAge  (years) | *Continuous (g/kg/day)* |  | -6.147 | -6.723, -5.571 | <0.0001 | -5.894 | -6.492, -5.295 | <0.0001 |
|  | *Q5 (>1.40)* |  | -8.039 | -8.789, -7.290 | <0.0001 | -7.378 | -8.150, -6.606 | <0.0001 |
|  | *Q4 (1.07-1.40)* |  | -5.773 | -6.879, -4.668 | <0.0001 | -5.361 | -6.395, -4.328 | <0.0001 |
|  | *Q3 (0.85-1.07)* |  | -3.489 | -4.239, -2.739 | <0.0001 | -3.170 | -3.901, -2.439 | <0.0001 |
|  | *Q2 (0.62-0.85)* |  | -2.149 | -2.987, -1.312 | <0.0001 | -1.948 | -2.767, -1.130 | <0.0001 |
| KDM  (years) | *Continuous (g/kg/day)* |  | -3.879 | -4.687, -3.070 | <0.0001 | -4.548 | -5.553, -3.542 | <0.0001 |
|  | *Q5 (>1.40)* |  | -4.927 | -5.971, -3.883 | <0.0001 | -5.672 | -7.019, -4.326 | <0.0001 |
|  | *Q4 (1.07-1.40)* |  | -3.293 | -4.461, -2.126 | <0.0001 | -3.771 | -4.993, -2.549 | <0.0001 |
|  | *Q3 (0.85-1.07)* |  | -1.894 | -2.825, -0.964 | 0.001 | -2.269 | -3.329, -1.210 | 0.001 |
|  | *Q2 (0.62-0.85)* |  | – | – | ns | – | – | ns |

Footnote: Values represent survey-weighted linear regression coefficients (β), 95% confidence intervals (95% CI), and p-values. Model 1 is adjusted for age (residual), sex, energy from carbohydrates (kcal/day), ethnicity, poverty-to-income ratio, tobacco use, physical activity, hypertension, and diabetes. Model 2 is adjusted for age (residual), sex, energy from fat (kcal/day), ethnicity, poverty-to-income ratio, tobacco use, physical activity, hypertension, and diabetes. All models account for the complex sampling design of NHANES using sample weights, strata, and primary sampling units. Reference group for quintiles: Quintile 1 (lowest protein intake). 'ns' denotes non-significant associations.

Supplementary Table 8*:* Association of protein intake and biological aging markers with cause-specific mortality in U.S. adults.

|  |  | **Heart diseases** | | |  | **Cerebrovascular** | | |  | **Malignant neoplasms** | | |
| --- | --- | --- | --- | --- | --- | --- | --- | --- | --- | --- | --- | --- |
|  |  | *Unadjusted HR (95%CI)* |  | *Adjusted HR (95%CI)* |  | *Unadjusted HR (95%CI)* |  | *Adjusted HR (95%CI)* |  | *Unadjusted HR (95%CI)* |  | *Adjusted HR (95%CI)* |
| **Protein intake** |  | 0.46 (0.31–0.67)* |  | 0.42 (0.16–1.11) |  | 1.11 (0.29–4.23) |  | 1.06 (0.09–11.96) |  | 0.52 (0.36–0.73)* |  | 0.36 (0.15–0.86)* |
| **Telomere length** |  | 0.05 (0.01–0.15)* |  | 0.32 (0.10–1.02) |  | 0.11 (0.03–0.47)* |  | 1.20 (0.23–6.28) |  | 0.05 (0.02–0.12)* |  | 0.28 (0.11–0.73)* |
| *Protein intake*  *+*  *Telomere length* |  | 0.50 (0.33–0.75)* |  | 0.45 (0.17–1.20) |  | 1.17 (0.30–4.60) |  | 1.05 (0.09–12.09) |  | 0.56 (0.39–0.81)* |  | 0.38 (0.16–0.91)* |
|  |  | 0.05 (0.02–0.17)* |  | 0.36 (0.12–1.10) |  | 0.11 (0.02–0.51)* |  | 1.19 (0.21–6.85) |  | 0.06 (0.02–0.14)* |  | 0.34 (0.13–0.85)* |
| **AnthropoAge** |  | 1.10 (1.08–1.13)* |  | 1.05 (1.00–1.10)* |  | 1.12 (1.03–1.22)* |  | 1.02 (0.80–1.30) |  | 1.08 (1.06–1.09)* |  | 1.06 (1.01–1.11)* |
| Protein intake  +  AnthropoAge |  | 0.64 (0.39–1.03) |  | 0.48 (0.16–1.40) |  | 2.08 (0.47–9.23) |  | 1.28 (0.14–11.63) |  | 0.70 (0.46–1.05) |  | 0.43 (0.16–1.12) |
|  |  | 1.10 (1.08–1.13)* |  | 1.02 (0.96–1.08) |  | 1.13 (1.05–1.21)* |  | 1.03 (0.82–1.29) |  | 1.08 (1.06–1.09)* |  | 1.03 (0.98–1.08) |
| **PhenoAge** |  | 1.11 (1.09–1.13)* |  | 1.05 (1.01–1.09)* |  | 1.11 (1.05–1.17)* |  | 1.01 (0.92–1.10) |  | 1.08 (1.06–1.09)* |  | 1.07 (1.02–1.12)* |
| *Protein intake*  *+*  *PhenoAge* |  | 0.64 (0.39–1.04) |  | 0.56 (0.24–1.29) |  | 2.21 (0.46–10.53) |  | 1.11 (0.10–12.14) |  | 0.71 (0.45–1.12) |  | 0.51 (0.22–1.19) |
|  |  | 1.11 (1.09–1.12)* |  | 1.04 (1.00–1.08)* |  | 1.12 (1.07–1.17)* |  | 1.01 (0.94–1.08) |  | 1.08 (1.06–1.09)* |  | 1.05 (1.00–1.10)* |
| **KDM** |  | 1.11 (1.09–1.12)* |  | 1.04 (1.00–1.08)* |  | 1.10 (1.03–1.17)* |  | 1.03 (0.96–1.11) |  | 1.07 (1.05–1.09)* |  | 1.03 (1.00–1.06) |
| *Protein intake*  *+*  *KDM* |  | 0.68 (0.44–1.06) |  | 0.51 (0.21–1.23) |  | 1.93 (0.55–6.81) |  | 1.19 (0.10–13.89) |  | 0.68 (0.45–1.02) |  | 0.39 (0.17–0.90)* |
|  |  | 1.10 (1.09–1.12)* |  | 1.04 (1.00–1.07)* |  | 1.11 (1.05–1.17)* |  | 1.03 (0.96–1.10) |  | 1.07 (1.05–1.09)* |  | 1.02 (0.99–1.04) |

Values are hazard ratios (HR) with 95% confidence intervals (95% CI) derived from Cox proportional hazards models. Unadjusted models include protein intake and each biological aging marker separately or jointly. Adjusted models were controlled for age (residual method), sex, total energy intake (kcal), race/ethnicity, poverty-to-income ratio, current smoking, physical activity, hypertension, and diabetes. All analyses accounted for the complex survey design of NHANES, including sampling weights, strata, and primary sampling units. Asterisks (*) indicate p < 0.05..

*Abbreviations*: HR, hazard ratio; CI, confidence interval; KDM, Klemera and Doubal’s method; PhenoAge, phenotypic age; AnthropoAge, anthropometric age.
